# A glomerular transcriptomic landscape of *APOL1* in Black patients with focal segmental glomerulosclerosis

**DOI:** 10.1101/2021.02.18.21251945

**Authors:** Michelle M. McNulty, Damian Fermin, Felix Eichinger, Dongkeun Jang, Matthias Kretzler, Noel Burtt, Martin R. Pollak, Jason Flannick, David J. Friedman, Nephrotic Syndrome Study Network (NEPTUNE), Matthew G. Sampson

## Abstract

Apolipoprotein L1 (*APOL1*)-associated focal segmental glomerulosclerosis (FSGS) is the dominant form of FSGS in Black people. There are no targeted therapies for this condition, in part because the molecular mechanisms underlying *APOL1’s* pathogenic contribution to FSGS are incompletely understood. Studying the transcriptomic landscape of *APOL1* FSGS in patient kidneys is an important way to discover genes and molecular behaviors that are unique or most relevant to the human disease. With the hypothesis that the pathology driven by the high-risk (HR) *APOL1* genotype is reflected in alteration of gene expression across the glomerular transcriptome, we compared expression and co-expression profiles of 15,703 genes in 16 Black FSGS patients with a HR vs 14 with a low-risk (“LR”) *APOL1* genotype. Expression data from *APOL1*-inducible HEK293 cells and normal human glomeruli were used to pursue genes and molecular pathways illuminated in these studies.

We discovered (1) increased expression of APOL1 in HR and nine other significant differentially expressed genes, including stanniocalcin (*STC1)*, which has a role in mitochondrial and calcium-related processes, (2) differential correlations between HR and LR *APOL1* and metabolism pathway genes, but similar correlations with extracellular matrix- and immune-related genes, (3) significant loss of co-expression of mitochondrial genes in HR FSGS, and (4) an NF-κB -down-regulating gene, *NKIRAS1*, as the most significant hub gene with strong differential correlations with *NDUF* family and immune-related genes. Overall, differences in mitochondrial gene regulation appear to underlie many differences observed between HR and LR FSGS. All data are available for secondary analysis through the “APOL1 Portal” (http://APOL1portal.org).

## Introduction

Positive selection events can result in common, population-specific variants. This is the case for the two exonic variants of apolipoprotein L1 (*APOL1*), G1 -comprised of two missense variants, and G2, a six base-pair deletion^1^. One copy of risk variant (RV) *APOL1* protects against trypanosomiasis. Two copies (a high-risk [“HR”] genotype) are associated with increased risk of multiple kidney diseases, including focal segmental glomerulosclerosis (FSGS) (17x increased odds)^1, 2 3^. 13% of Black Americans carry the HR *APOL1* genotype, but this rises to 60-70% in those with FSGS^3, 4^. These RVs are rare in individuals without recent African ancestry^5^. Thus, the added kidney disease risk from *APOL1* uniquely affects Black individuals, who already face disproportionate health burdens due to structural racism and other negative social drivers^6^.

To ultimately develop treatments and cures for *APOL1*-associated FSGS, we must discover the mechanisms by which its RVs cause/contribute to it. While *APOL1* is absent outside of humans and some primates, transgenic animals and cell lines have provided insights into disease pathogenesis^7-9^. Insights include; (1) increased *APOL1* expression by immune activation^10^ (2) toxicity associated with the RV and increased expression^8, 11^, (3) *APOL1* ion channel formation and function dependent on complex intracellular trafficking and pH changes^12, 13^, and (4) multiple pathways implicated in cellular damage including mitochondrial dysfunction^14-16^, inflammasome activation^17^, ER stress^18, 19^, increased cation transport^13, 20^, and global protein synthesis inhibition^21,22^.

However, a gap remains in understanding the molecular components driving the *APOL1*-associated FSGS phenotype in patients. Dissecting the transcriptomic landscape of kidney tissue from affected patients – where *APOL1*’s regulation and expression are within its native genomic context – can help address this gap. *APOL1*-associated FSGS genes, pathways, or transcriptional behaviors discovered this way would arguably have increased specificity and relevancy towards understanding the human disease.

Our conceptual model of *APOL1’s* role in FSGS was: (1) a HR genotype primarily exerts its effects via a change in *APOL1* function (given that the RV are protein-altering variants) rather than massive changes in its own expression and (2) this HR-specific *APOL1* function results in differences in cellular programs which are potentially revealed through changes in gene expression and co-expression of glomerular genes. To test this model, we performed glomerular transcriptome-wide differential expression and differential co-expression studies comparing Black patients with biopsy-proven FSGS^23^ with a low-risk (“LR”; 0 or 1 risk alleles) or HR genotype (**Figure 1**).

**Figure 1.**
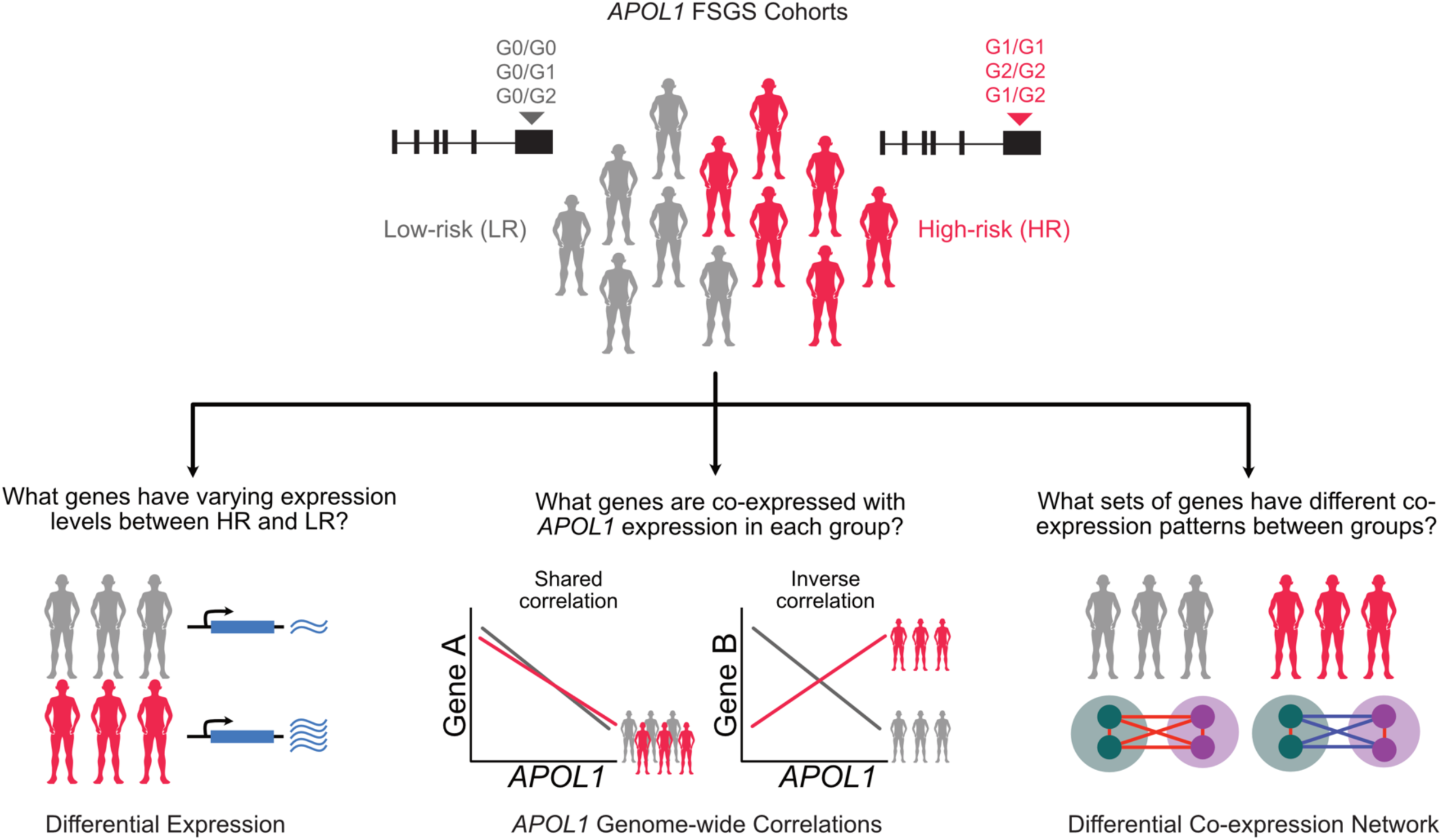
Analysis schematic: *APOL1* risk variants were genotyped for Black participants with FSGS from the NEPTUNE cohort. There were 16 high-risk (“HR,” 2 risk variants) and 14 low-risk (“LR,” 0 or 1 risk variant) participants. Three glomerular transcriptomic analyses were used to illuminate mRNA expression differences as a function of risk genotype and *APOL1* expression. (1) **differential expression** to identify genes with varying expression between the HR and LR FSGS group. (2) **transcriptome-wide correlation of single genes with *APOL1*** to identify similarities and differences between genes coexpressed with *APOL1* specifically in HR versus LR state. (3) **differential co-expression** to go beyond solely *APOL1* correlations and identify groups of genes whose co-expression differs with each other in the HR and LR *APOL1* state. The latter two analyses empowered identification of potential molecular perturbations between groups that are not reflected through differential expression.

## Methods

The work described has been carried out in accordance with The Code of Ethics of the World Medical Association (Declaration of Helsinki).

### NEPTUNE

Samples were from the Nephrotic Syndrome Study Network (NEPTUNE)^23^, a biopsy-based, longitudinal study of patients with proteinuric kidney diseases. We selected FSGS patients with a self-reported race/ancestry ‘Black/African American’ or a genotype-based African ancestry from Peddy^24^.

### APOL1 Genotyping

Twenty-four samples were genotyped through previously described whole-genome sequencing (WGS) and Sanger sequencing^25, 26^, five through WGS alone, and one using RNA-seq.

### Glomerular RNA-seq

Using standard methods, glomerular mRNA underwent sequencing and quality control followed by alignment, gene expression counting of protein-coding genes, and normalization^27-32^ (see **Supplement** for details).

### Cell-Type-Specific Gene Expression

Using the genes unique to a cell type^33^, we approximated the cell-type abundances as the mean of the cell-type-specific quantile normalized gene expression in each sample. The difference in mean cell-type expression between groups was tested with a Wilcoxon test and a Bonferroni adjusted significance threshold of 0.05/24=0.002.

### Differential Expression

We conducted the differential expression analysis with DESeq2^30^ using the filtered gene count matrix from StringTie, adjusting for age, sex, and RNAseq batch. Cook’s distance was used to flag associations influenced by outliers. Results were plotted with EnhancedVolcano [https://github.com/kevinblighe/EnhancedVolcano].

### *APOL1* Genome-Wide Correlations

In each group, we calculated Spearman correlations between *APOL1* and normalized expression for each gene. The ranked correlations were assessed by Webgestalt (WEB-based Gene SeT AnaLysis Toolkit) ^34^ for analysis of enrichment of “annotation terms”. Enriched annotation terms with FDR < 0.05 in both HR and LR analyses were considered overlapping; the sign of the normalized enrichment score was used to determine if the enrichments were concordant or discordant. A threshold of FDR < 0.2 was used to compare enrichments with normal tissue.

### DiffCoEx

Differential co-expression gene modules were built using methods outlined by DiffCoEx^35^ (see Supplement for full details). Briefly, we calculated the adjacency matrix of Spearman correlations for HR and LR separately and computed the matrix of adjacency differences, ranging from 0 to 1 (where 1 indicates the strongest difference in correlations). This created a topology overlap matrix which was then clustered into modules with Weighted Gene Correlation Network Analysis (WGCNA). Intra-modular connectivity for each gene was defined as the sum of adjacencies (from the matrix of adjacency differences) for each gene normalized by the largest connectivity value. Hub genes were defined as genes with intra-modular connectivity greater than 0.7. Submodules were identified by comparing the genes’ hierarchical clustering within a DiffCoEx module in HR and LR separately. Submodule eigengenes (1^st^ principal component) were used to compare patterns between submodules. We used Cytoscape (v3.7.2) to visualize the correlation networks.

### Enrichment Analyses – Webgestalt, DAVID

We used Webgestalt for gene set enrichment analyses. Correlations were ranked by decreasing magnitude in each group separately. We tested enrichment in all Gene Ontology (GO) categories, KEGG, Panther, Wikipathway, and Reactome databases. 2,000 simulations were used to generate p-values. FDR was from reference dataset-specific adjustments. DAVID (v6.8)^36, 37^ was used for over-representation analyses of gene modules from DiffCoEx, using all our analysis genes as background.

### CMAP

To test enrichment in the Broad Connectivity Map (CMAP)^38^, we selected genes with |log2 fold change| >0.75 and an unadjusted p-value < 0.05, excluding genes flagged by Cook’s distance or low counts, and reported results from their HA1E cell-line only. CMAP connectivity scores, ranging from −100 to 100, quantify the relationship between the query signatures and our differentially expressed genes. A score of 95 indicates that only 5% of the reference datasets have shown stronger connectivity than the query.

### HEK293 Analyses

As previously published^20^, HEK293 cells were stably transfected with *APOL1* on a TET ON promoter and harvested nine hours after tetracycline induction. DESeq2 was used to test for differential expression between four G1 replicates and two G0 replicates. Genes included were those with greater than ten normalized counts in all samples. Cook’s distance filtered was not applied due to the small sample size. Only replicated genes are reported.

### Statistical Analyses

R Studio Version 1.2.5033 was used for all analyses and figures created with ggplot, ggcorplot, and ggpairs.

## Results

Participants were from NEPTUNE^23^, a biopsy-based cohort of patients with proteinuric kidney diseases. Inclusion criteria for this study were a histologic diagnosis of FSGS, glomerular RNA-seq data, *APOL1* genotyping, and self-identified ‘Black/African-American’ race or genotype-predicted African continental ancestry. This resulted in 14 LR and 16 HR samples.

There were no significant demographic or clinical differences at the time of biopsy (**Table S1, Table 1**). There were no significant differences in cell-type distribution between HR and LR groups based on a comparison of the mean expression of cell type-specific genes^33^ in the glomerular transcriptome data (**Figure S1**). These data support the validity of comparing these two groups as a function of *APOL1* genotype with limited confounding by phenotypic or cell type differences.

**Table 1:** Participant Demographics

|  | Low-Risk | High-Risk | P<sup>a</sup> |
| --- | --- | --- | --- |
| n | 14 | 16 |  |
| Sex = Male (%) | 9 ( 64.3) | 9 ( 56.2) | 0.72 |
| Age of Biopsy (median [IQR]) | 29.5 [13.3, 57.0] | 16.5 [15.8, 26.8] | 0.38 |
| Age of Onset (median [IQR]) | 20.0 [4.8, 56.3] | 16.0 [15.0, 23.0] | 0.98 |
| Race (%) |  |  | 0.49 |
| Black/African American | 13 ( 92.9) | 13 ( 81.2) |  |
| Multi-Racial | 1 ( 7.1) | 0 ( 0.0) |  |
| White/Caucasian | 0 ( 0.0) | 1 ( 6.2) |  |
| Unknown | 0 ( 0.0) | 2 ( 12.5) |  |
| Black (%) |  |  | 0.49 |
| Yes | 14 (100.0) | 13 ( 81.2) |  |
| No | 0 ( 0.0) | 2 ( 12.5) |  |
| NA | 0 ( 0.0) | 1 ( 6.2) |  |
| Genotype-predicted Ancestry (%) |  |  | 0.49 |
| African | 13 ( 92.9) | 13 ( 81.2) |  |
| Admixed American | 0 ( 0.0) | 2 ( 12.5) |  |
| Unknown | 1 ( 7.1) | 0 ( 0.0) |  |
| NA | 0 ( 0.0) | 1 ( 6.2) |  |
| Immunosuppression at Biopsy = yes (%) | 5 ( 35.7) | 4 ( 25.0) | 0.69 |
| eGFR at Biopsy (median [IQR]) | 66.7 [47.7, 89.4] | 62.9 [45.9, 71.1] | 0.28 |
| UPCR at Biopsy (median [IQR]) | 2.8 [1.4, 5.5] | 2.3 [1.1, 6.1] | 0.90 |
| *APOL1* Risk Alleles (%) |  |  | <0.01 |
| 0 | 3 ( 21.4) | 0 ( 0.0) |  |
| 1 | 11 ( 78.6) | 0 ( 0.0) |  |
| 2 | 0 ( 0.0) | 16 (100.0) |  |
| *APOL1* Haplotype (%) |  |  | <0.01 |
| G0_G0 | 3 ( 21.4) | 0 ( 0.0) |  |
| G0_G1 | 6 ( 42.9) | 0 ( 0.0) |  |
| G0_G2 | 5 ( 35.7) | 0 ( 0.0) |  |
| G1_G1 | 0 ( 0.0) | 5 ( 31.2) |  |
| G1_G2 | 0 ( 0.0) | 7 ( 43.8) |  |
| G2_G2 | 0 ( 0.0) | 4 ( 25.0) |  |

<sup>a</sup> Continuous variables were tested with non-parametric Wilcoxon rank-sum tests, categorical variables were tested with Exact tests. IQR= interquartile range, eGFR = estimated glomerular filtration rate, UPCR = urine protein/creatinine ratio,

### Differential expression identified increased HR *APOL1* expression and nine genome-wide significant genes

Here, in contrast to a previous study that included non-FSGS patients and used array-based expression^39^, glomerular expression of *APOL1* was significantly higher in the HR state (log_2_FC =0.57, p=0.02, **Figure 2**). Among 15,703 genes, we discovered nine genes with significant differential glomerular expression (FDR < 0.05) between HR and LR FSGS (**Table 2, Figure 2, Table S2)**. Stanniocalcin 1 (*STC1*) had the largest up-regulation (log_2_FC = 3.50, p_adj_=0.02) in HR, and C-C motif chemokine ligand 18 (*CCL18*) had the largest down-regulation (log_2_FC = −4.94, p_adj_=0.02).

**Table 2:** Differentially Expressed Genes

| Gene | Log2 FC | P-value | P-adj |
| --- | --- | --- | --- |
| *STC1* | 3.50 | $8.07 \times 10^{-6}$ | 0.02 |
| *CCN3* | 1.89 | $6.63 \times 10^{-6}$ | 0.02 |
| *CCN6* | 1.42 | $7.83 \times 10^{-6}$ | 0.02 |
| *HIPK1* | 1.23 | $2.21 \times 10^{-6}$ | 0.01 |
| *ANO5* | 1.07 | $1.96 \times 10^{-6}$ | 0.01 |
| *ALDOC* | 0.72 | $1.42 \times 10^{-5}$ | 0.02 |
| *DGKD* | -0.90 | $5.84 \times 10^{-7}$ | 0.01 |
| *NOS2* | -1.76 | $2.40 \times 10^{-5}$ | 0.04 |
| *CCL18* | -4.94 | $8.40 \times 10^{-6}$ | 0.02 |

Log2 FC = log2 fold change, positive values are upregulated in HR, compared to LR. P-adj = FDR adjusted p-value

**Figure 2.**
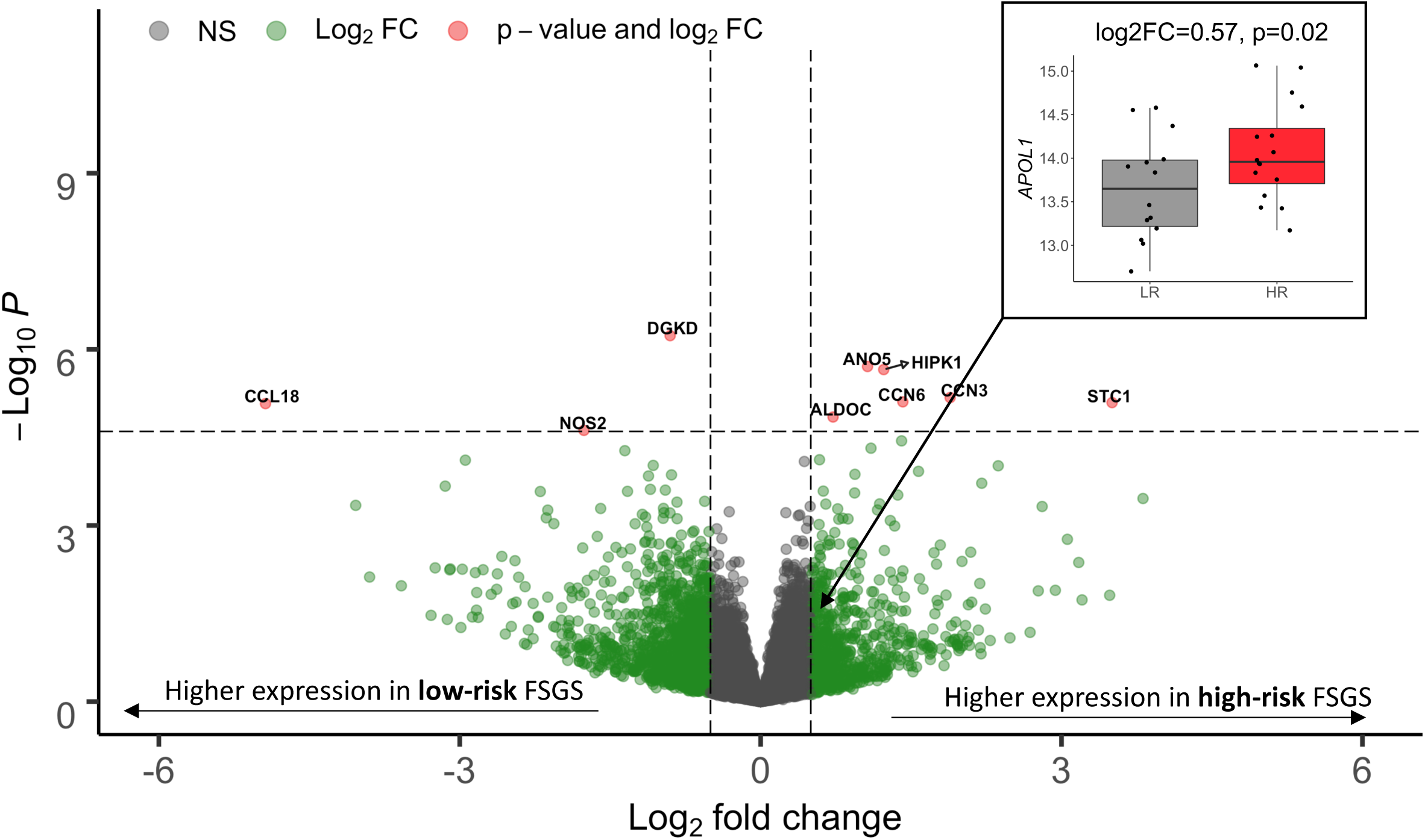
Differential expression volcano plot: Genes (represented as dots) up-regulated in high-risk samples, as compared to low-risk samples, are indicated with positive log2 fold change. Vertical hashed bars indicate log2 fold change of −0.5 and 0.5, which we define as the threshold for differential expression. The horizontal hashed bar indicates the adjusted significant p-value threshold 0.05 (P_unadjusted_ ∼ 2.5×10-5). Red dots identify the nine genes that passed the multiple testing correction with absolute fold change > 0.5. Green dots are differentially expressed genes that do not pass our significance threshold. Insert - Differential expression of *APOL1* in the low risk (gray) and high risk (red) states.

We followed-up this result using an independent dataset of HEK293 cell gene expression data measured after induction of either wild-type (G0) or G1 *APOL1* transgenes^20^. Four of the six up-regulated genes (*STC1, ALDOC, CCN3, HIPK1*) replicated differential expression in this system (p_adj_ < 0.05; **Table S3**). These *in vitro* results from non-immune cells suggest that increased HR *APOL1* expression in FSGS patients may be causally related to these genes’ glomerular expression changes and that their expression in FSGS patients does not solely come from non-resident immune/inflammatory cells.

### Connectivity Map highlights the role of calcium channels upstream differential expression

To infer potential drivers of the differential expression signature observed between HR and LR FSGS, we used an *in silico* method, the Connectivity Map (CMAP)^38, 40^. CMAP is a catalog of gene expression signatures of cell lines after being directly perturbed via chemical administration, gene knockdown, or gene overexpression. The CMAP database includes an immortalized human kidney epithelial cell line (HA1E). We hypothesized that an HAE1 differential expression signature observed between unexposed cells and those exposed to a subset of chemical or transcriptomic perturbations would either match or be the opposite of the differential expression signature in our FSGS samples (comprised of 235 genes with an |FC| > 0.75 between HR and LR and p_unadj_ < 0.05).

The expression signatures for the top perturbagens that match or oppose the *APOL1* FSGS signature are presented in **Figure 3**. For chemicals, a strong positive match was synephrine (connectivity score=99.61), an adrenergic receptor agonist and L-type calcium channel activator; and a strong opposing match was levetiracetam (connectivity score=-99.23), an N-type calcium channel inhibitor. Together, these results support a difference in calcium channel regulation between HR and LR FSGS patients that is contributing to the differentially expressed glomerular genes, with higher calcium channel activation in HR patients. Full CMAP results are in **Table S4**.

**Figure 3.**
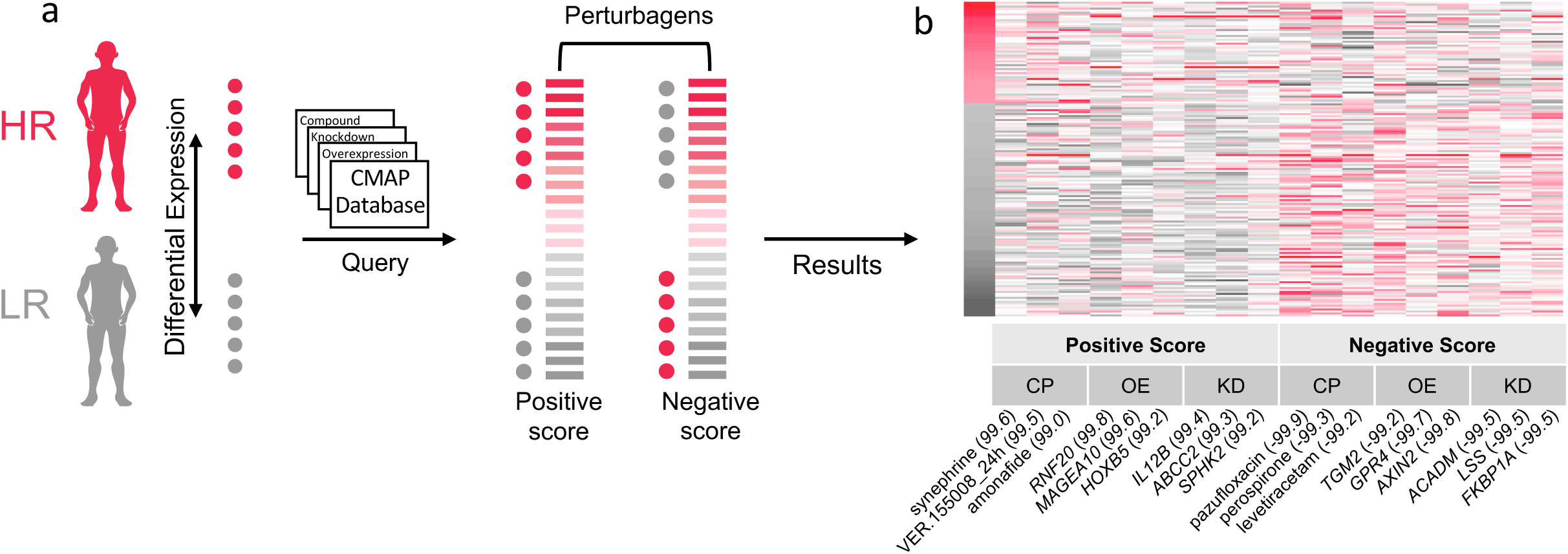
Connectivity Map (CMAP): **A)** Schematic of CMAP analysis. Differentially expressed glomerular genes in the high risk (HR) versus low risk (LR) state with absolute log2 fold change > 0.75 and unadjusted p-value < 0.05 were selected for the CMAP query. Red dots indicate genes with higher expression in the HR group, and gray dots indicate genes with higher expression in LR group. This gene set pattern was then compared to CMAP analyses of the impact of perturbagens - including compounds, gene knockdowns, and gene overexpression – on gene expression of the HAE1 kidney cell. Perturbagens with a positive score match our analysis, i.e., genes increased/decreased in HR are also increased/decreased in response to the perturbagen. Perturbagens with a negative score inversely match our analysis, i.e., genes increased/decreased in HR are decreased/increased in response to the perturbagen. **B)** Heatmap of CMAP HAE1 z-scores from the top three positive and negatively enriched perturbations for each category (KD=knockdown, OE=overexpression, CP=compound). Genes (y-axis) are sorted by the log2 fold change in the high-risk versus low-risk FSGS differential expression analysis. Numbers in parentheses indicate percentile of match among all perturbagens applied to HAE1 cells.

### Gene set enrichment analyses of *APOL1* correlations highlight enriched pathways unique to and shared by high-risk and low-risk groups

We then aimed to test our model that the HR genotype substantially alters *APOL1’*s function, leading to altered co-expression with other glomerular genes and ultimately to differences in cellular biology between risk states. To do this, we first computed expression correlations of 15,703 genes with *APOL1* (HR vs LR separately), ranked them by correlation magnitude, and performed gene set enrichment analysis(GSEA)^34^ to identify biologic pathways that were shared and different between the *APOL1* risk states (**Tables S5, S6**).

First, most of the significantly enriched pathways (FDR < 0.05) were unique to LR and HR - 94% and 87%, respectively. The vast majority of HR enriched pathways comprised genes positively correlated with *APOL1*, whereas enriched pathways in the LR group derived from positively and negatively correlated groups of genes (**Figure 4A**).

**Figure 4.**
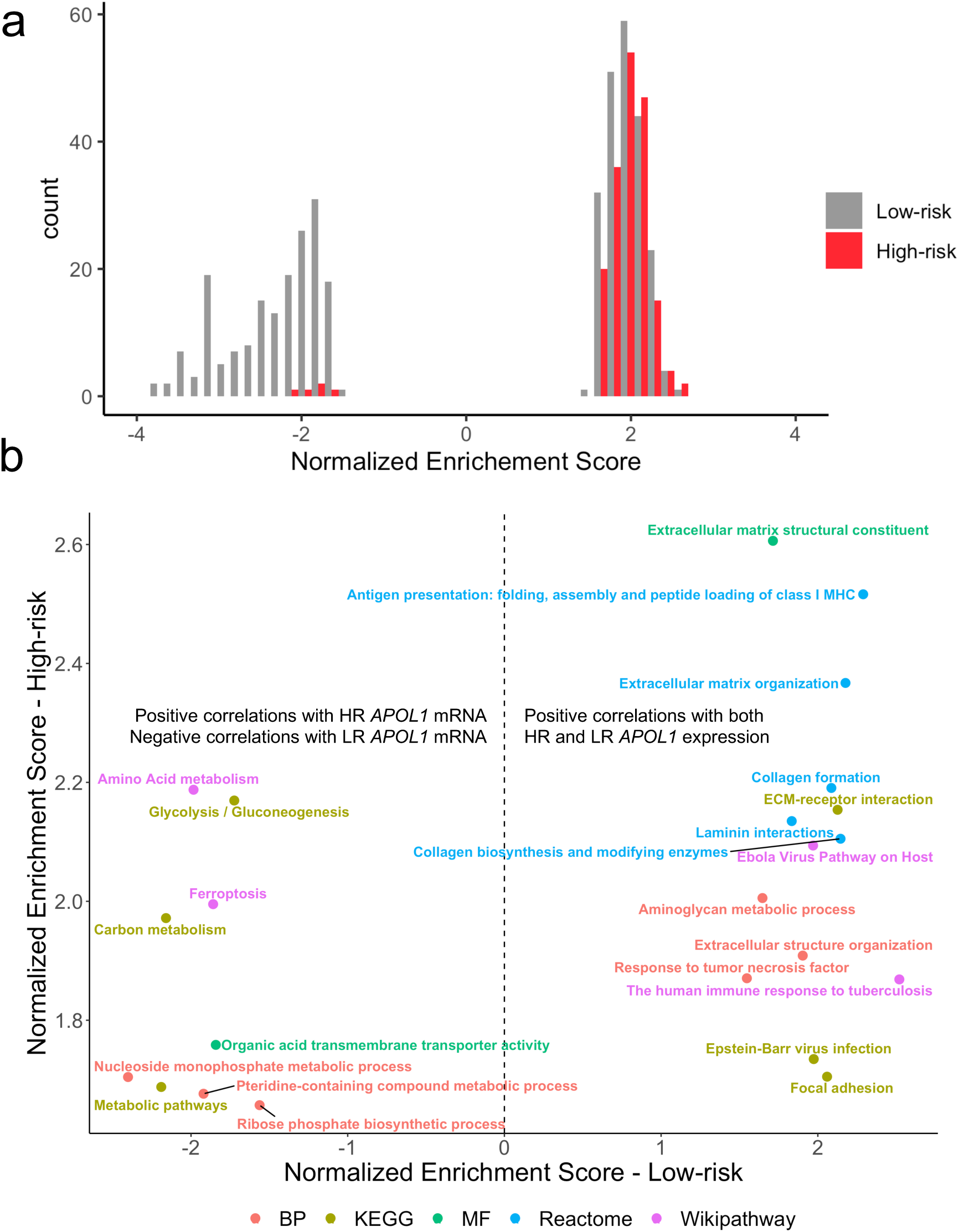
*APOL1* genome-wide correlations: **A)** Histogram of gene set enrichment analysis (GSEA) normalized enrichment scores (NES) with FDR < 0.05, stratified by risk genotype. Enrichments among genes negatively correlated with *APOL1* are indicated by a negative NES, and enrichments among genes positively correlated with *APOL1* are indicated by a positive NES. In the low risk (LR) group, there was enrichment among genes positively and negatively correlated with *APOL1*. In high risk (HR), most enrichments were among genes positively correlated with *APOL1* expression. **B)**Comparison of enrichment terms common to both HR and LR FSGS colored by gene annotation dataset. Annotations to the right of the hashed line indicate gene sets enriched among genes positively correlated with both HR and LR *APOL1* expression. Annotations to the left of the hashed line indicate gene sets negatively correlated with LR *APOL1* expression and positively correlated with HR *APOL1* expression.

Twenty-three pathways were significantly correlated with both HR and LR *APOL1* expression (FDR < 0.05). Fourteen of these were comprised of genes positively correlated with both HR and LR APOL1 expression. These included gene sets related to the extracellular matrix (e.g., collagen formation, laminin interactions) and the immune system (**Figure 4B**). The role of *APOL1* in innate immunity has been established^10^. However, the shared positive correlation of HR and LR *APOL1* with extracellular matrix genes (e.g., laminins, integrins, collagen) in FSGS has not been reported in either risk state. These results suggest that targeted study of *APOL1’s* potential coregulation and/or biological interactions with the extracellular matrix and these specific genes could be fruitful for further mechanistic understanding (**Data File S1**).

The component genes of 9 of 23 gene sets were significantly correlated in a positive direction with HR *APOL1* but negatively correlated with LR *APOL1*; many of them related to metabolism (**Figure 4B**). The strongest of these discordant annotations were the Wikipathways term “Amino Acid Metabolism” and the KEGG Pathway “Glycolysis/Gluconeogenesis.” An additional discordant term was “Ferroptosis,” a form of programmed cell death caused by iron-dependent phospholipid peroxidation^41^. In HR *APOL1* glomeruli of FSGS patients, expression of genes in the ferroptosis pathway increase as well. Autophagy has been implicated as one potential mechanism participating in *APOL1* associated glomerular disease^11, 42, 43^, and the *APOL1* family of proteins, in general, are known to function in cell death pathways^44, 45^. The extent to which any of these pathways has a role in *APOL1* kidney disease requires further experimental clarification.

To triangulate which *APOL1* correlation signature may be aberrant, we compared *APOL1* correlation enrichment of the nine discordantly-correlated pathways from normal-appearing glomeruli microdissected from nephrectomies. With this data, we could more confidently assign the “disease state” to the FSGS subtype whose signature was discordant with the normal glomerular expression signature. In the normal tissue, there was an enrichment of three of these pathways (FDR< 0.2); “pteridine-containing compound,” “nucleoside monophosphate,” and “organic acid transporter activity” (**Table S7**). In all three, the patterns in the normal tissue matched LR FSGS. This suggests that for these pathways, HR *APOL1’s* correlation patterns with its co-expression partners represent the pathologic state.

### Differential co-expression identifies gene modules and submodules with changes in correlation patterns between *APOL1* states

Independent studies have demonstrated that differentially co-expressed genes are often not differentially expressed^46^. Furthermore, even without differential expression, groups of genes comprising regulatory networks can differentially co-express (“re-wire”) between biologic conditions^47^. As such, we hypothesized that biological pathways disrupted in *APOL1*-associated FSGS might be reflected by these types of changes in transcriptomic network relationships and not limited to differentially expressed genes or those only correlated with *APOL1*. To test this, we turned to the differential co-expression method^35^ (“DiffCoEx”). Using the same 30 samples and 15,703 genes from previous analyses, we used DiffCoEx to identify and cluster genes whose correlation patterns changed between HR and LR FSGS patients’ glomeruli.

1,273 genes showed varying degrees of differentially co-expression with each other, clustering into four modules (“Green”-137 genes, “Purple”-40 genes, “Pink” – 847 genes, “Brown” – 249 genes; **Figure 5A**). These modules were defined based on their component genes’ magnitudes and direction of correlation within themselves - and in relation to other modules - in the HR vs LR state. By clustering each module’s genes separately in the HR and LR state, we further refined them into smaller, putatively more specific, “submodules” (**Figure 5B, 5C**). Once the DiffCoEx modules and submodules were defined, we sought to more clearly understand relationships within and between them and to identify enriched gene sets and key genes – “hub genes” (**Figure 5D, Table S2, Figure S4, S5**).

**Figure 5.**
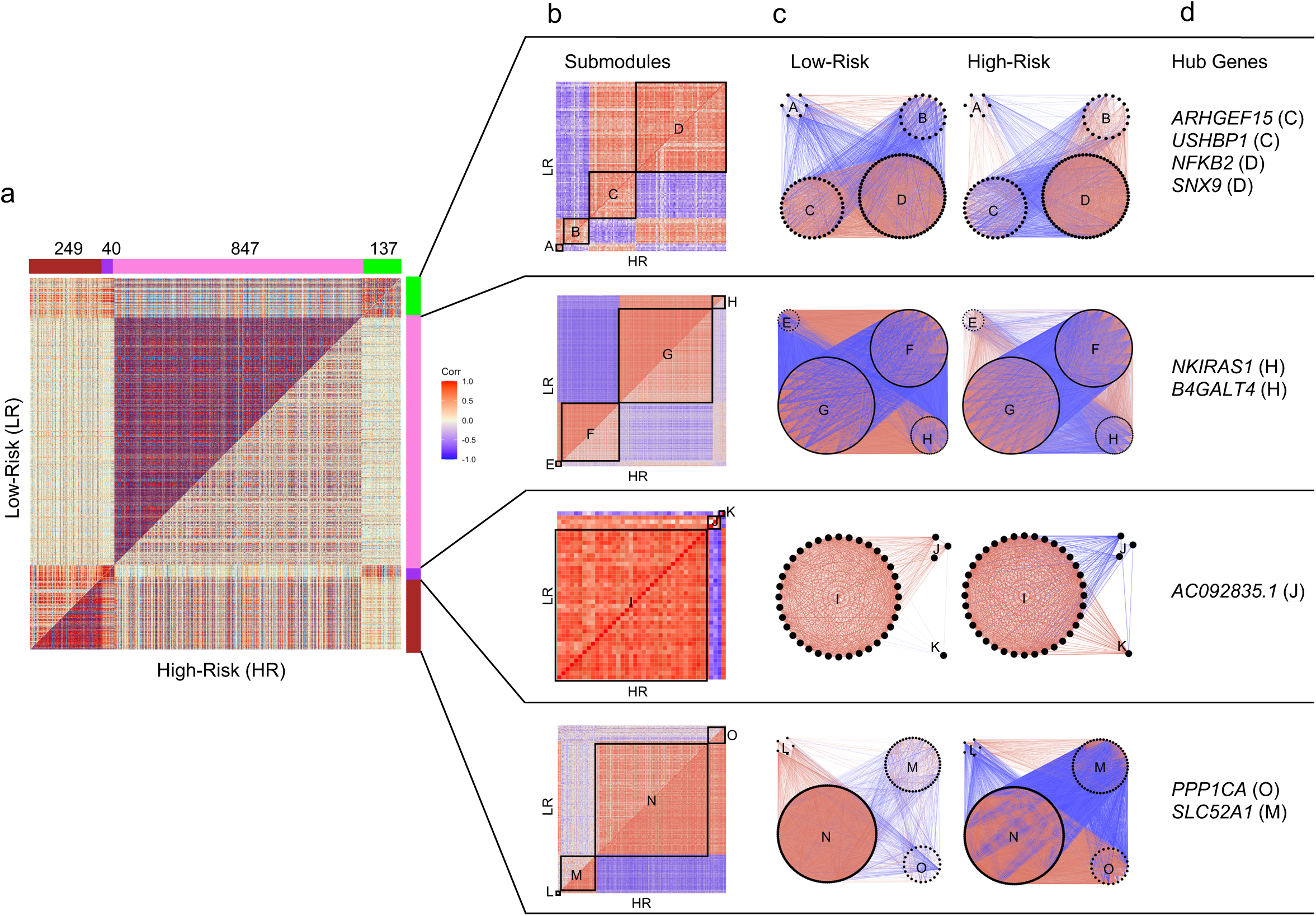
Differential co-expression analysis of 15,703 protein-coding genes in high-risk versus low-risk FSGS: (Positive correlations are red and negative are blue throughout all figures). **A)** Symmetric correlation heatmap of differentially co-expressed genes grouped into four modules (“Brown,” “Purple,” “Pink,” “Green”). Each row and column is a single gene. The upper triangle (above the diagonal) reflects correlations in the low risk (LR) samples, and the lower triangle (below the diagonal) the high risk (HR) samples. The number of genes in each module is indicated above the module color. **B)** Submodule correlation heatmaps for each differentially co-expressed gene module, identified by hierarchical clustering of module genes in each group. LR correlations are reflected on the upper triangle and HR on the bottom. Submodules are labeled alphabetically. **C)** Gene network view of correlations within and between gene submodules stratified by risk genotype. **D)** Hub genes (most differentially co-expressed genes), for each module with the corresponding submodule label. Genes with a normalized connectivity > 0.7 were considered hub genes.

### Disruption of co-expression in high-risk samples include genes integral to the mitochondrial inner membrane and transcription processes

The Pink module was defined by stronger co-expression of its 847 genes in the LR state, with 91% of gene pairs having a stronger absolute correlation in LR. There was significant enrichment for terms such as “mitochondrial inner membrane,” “oxidative phosphorylation,” “mitochondrial respiratory chain complex I,” “mRNA splicing,” and “ribonucleoprotein” (**Table S8**). Previous studies have implicated mitochondrial dysfunction, abnormal energetics and reduced transcription in *APOL1* associated FSGS^14-16, 21, 48, 49^. Given its large size and its enrichment for genes, subcellular structures, and pathways previously implicated in *APOL1* FSGS, we focused on more deeply characterizing the Pink module and its submodules. In particular, given the weaker co-expression in the HR state, we hypothesized that genes in the Pink module might harbor sets of genes with aberrant coregulation that could contribute to, or be the result of, the inciting pathobiology of *APOL1*-associated FSGS.

There were four Pink submodules (“Pink_E,” “Pink_F,” “Pink_G,” “Pink_H”). We performed an over-representation analysis of each submodule using DAVID (**Table S9**). We also computed eigengene correlations between them in the HR and LR FSGS state and in normal tissue (**Figure 6A**). Enriched terms in submodules Pink_E and Pink_F were UniProt terms “spliceosome” and “ATP binding,”and “nucleoplasm.” Genes in submodules Pink_G and Pink_H were mitochondrially enriched and included terms such as GO term “oxidative phosphorylation.” Notably, the Pink_H submodule was enriched for the mitochondrial inner membrane translocase subunit (TIM complex – *TIMM17A, TIMM23, TIMM23B*). A previous study found that the TIM complex was required for *APOL1* import into the mitochondrial matrix and was a precursor of *APOL1*-induced cytotoxicity^16^.

**Figure 6.**
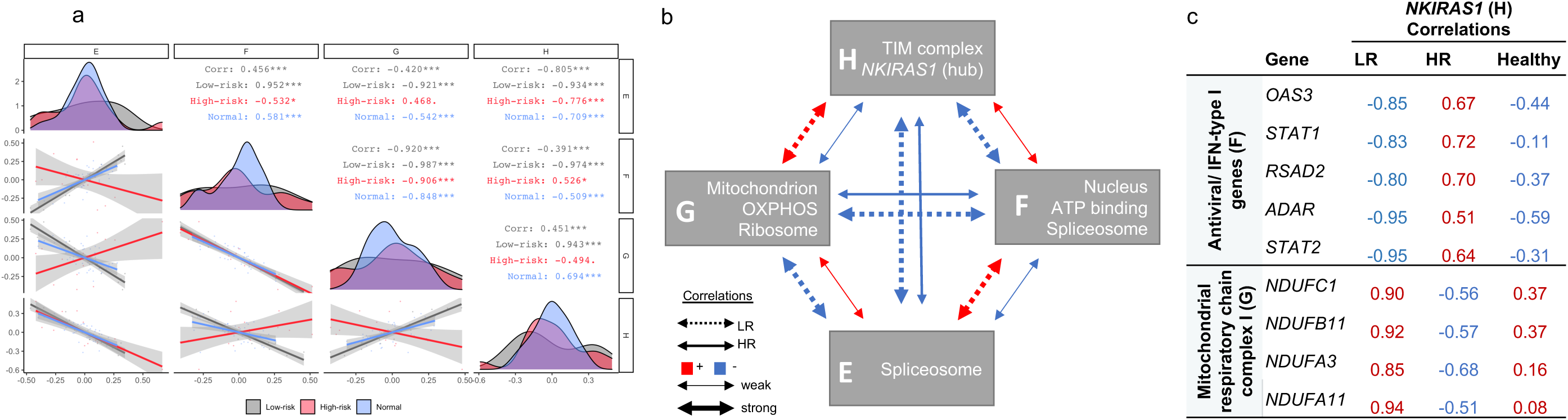
Characterization of the ‘Pink’ differentially co-expressed module: **A)** Spearman correlation of submodule eigengenes in high risk (HR), low risk (LR), and normal glomerular tissue (HR – red; LR – black, normal – blue). Eigengenes summarize overall gene expression behavior in each submodule and were defined as the first principal component of the group-specific gene expression matrices. Submodules are labeled along the x and y-axis. Diagonal – eigengene density plots. Below diagonal – scatter plot comparing submodule eigengenes (each point represents a sample). Above diagonal – Spearman correlations stratified by group (“Corr” – all groups combined). Correlation significance - “***” p < 0.01, “**” p<0.01, “*” p<0.05, “.” p<0.1. **B)** HR and LR correlation schematic of the four ‘Pink’ submodules with selected enriched biology. **C)** We identified the top 10% of Pink module genes differentially co-expression with NKIRAS1, resulting in 30 and 55 genes from F and G submodules, respectively. Presented here are HR, LR, and healthy NKIRAS1 correlations with other genes from the corresponding enrichment analysis. These genes are among the strongest differentially correlated genes with NKIRAS also enriched for known biology. Absolute correlations are stronger in the LR group compared to the HR group and correlation patterns in the healthy tissue are more similar to LR, compared to HR group.

We present a summary of the enriched biology and the overall co-expression patterns in the Pink submodules in **Figure 6B**. Overall there were opposite directions of co-expression between each submodule by risk genotype, and the magnitude of correlation was always stronger in the LR state. The normal tissue co-expression patterns were more similar to LR than HR (**Figure 6A**). From this, we deduce that the co-expression patterns of these glomerular genes in HR *APOL1* represent the diseased state.

### Differential co-expression of *NKIRAS1* highlights potential high-risk-specific gene regulation changes involving JAK/STAT and mitochondrial complex 1 genes

In differential co-expression networks, hub genes are those with the most substantial differences in correlations with other genes between two comparative groups^47^. Here, the strongest hub gene was NF-κB inhibitor interacting ras like 1 (*NKIRAS1)*, located within submodule Pink_H (**Table S2**). *NKIRAS1* down-regulates NF-κB by inhibiting the degradation of one of its inhibitors, NF-κB inhibitor beta (*NFKBIB*)^50 51^. In LR, *NKIRAS1* has strong positive correlations with genes in submodule Pink_G, a relationship that becomes non-significant or negative in HR. Conversely, *NKIRAS1* has strong negative correlations with submodule Pink_F in LR, a relationship that becomes non-significant or positive in HR

We hypothesized that genes showing the strongest differences in co-expression with *NKIRAS1* might illuminate regulatory pathways unique to, or dysregulated in, *APOL1*-associated FSGS. We focused our enrichment analysis on the top 10% of genes from the Pink module with the greatest difference in *NKIRAS1* correlations (**Tables S2, S10**). **Figure 6C** lists correlations between *NKIRAS1* and its top differentially co-expressed, biologically enriched genes. The Pink_F submodule genes had the strongest enrichment for the type I interferon signaling pathway (p_adj_=7.3×10^−4^), including *STAT2*, which can bind to *APOL1’s* transcription start site^10^. Of note, five of the 7 *JAK/STAT* genes were clustered to submodule Pink_F; comparison of *JAK/STAT* correlations with *NKIRAS1* are in **Figure S3**. The Pink_G module genes were enriched in the mitochondria (p_adj_=5.7 ×10^−3^), and more specifically, mitochondrial respiratory chain complex I, (p_adj_=2.1 ×10^−2^). This includes four NADH:ubiquinone oxidoreductase (NDUF) genes composing complex 1 of the respiratory chain. Given the stronger correlation strengths of *NKIRAS1* in the LR groups and shared correlation patterns between normal kidney and LR glomeruli, we hypothesize that a regulation pathway, with inverse regulation of immune and mitochondrial genes, is being disrupted in HR.

## Discussion

Given that the majority of Black patients with FSGS have *APOL1-*associated disease, developing treatments and cures targeting it can make a major health impact in this population. To this end, we used three complementary glomerular transcriptomic analyses to illuminate genes, pathways, and networks that may be contributing to, or driving, the pathobiology of *APOL1-*associated FSGS. We focused exclusively on Black FSGS patients to increase confidence that differences observed were more likely to be specific to *APOL1* risk genotype and not confounded by genetic ancestry, structural racism, or other NS histologies. And by comparing two groups of FSGS patients differing significantly only by *APOL1* status (rather than a case-control study), we could be more certain that changes we discovered were related to *APOL1*-associated FSGS rather than FSGS non-specifically.

A major distinction between HR and LR FSGS related to differences in expression and/or co-expression of genes localized to mitochondrial compartments and/or processes. In the past decade, many experimental studies using inducible cell lines have linked increased HR *APOL1* expression to mitochondrial dysfunction ^14-16, 21, 48, 49^. Here, we extend these *in vitro* findings to patients with FSGS, increasing the relevancy and importance of dissecting these pathways. Furthermore, there was a closer match of the mitochondrial co-expression signatures of normal tissue with the LR state. Thus, we are more confident in classifying the mitochondrial co-expression patterns in HR FSGS glomeruli as pathologic.

*STC1* was the most significantly increased gene in HR glomeruli. It is notable that in a previous genetic study of Black individuals with ESKD, a variant at the *STC1* locus had a weak statistical interaction with *APOL1*^52^. *STC1*, along with two of the other five most up-regulated genes - *HIPK1* and *CCN6* – have been shown to regulate mitochondrial function through cytoprotective, antioxidant pathways^53-55^. In mesenchymal stromal cells, the up-regulation of *STC1* uncouples oxidative phosphorylation, reduces reactive oxygen species, and switches metabolism towards glycolysis^53^. Supporting this, we discovered significant enrichment for glycolysis genes among positive *APOL1* correlations specifically in the HR state. We found that induction of the *APOL1* RV in the HEK293 cells resulted in increased *STC1* expression. Future work is needed to test the hypothesis that increased *STC1* expression is triggered to help rescue cells from *APOL1*-induced cytotoxicity.

Our differential co-expression data further supported disruption of mitochondrial function in HR FSGS, with co-expression changes between TIM complex genes *TIMM17A, TIMM23*, and *TIMM23B* and mitochondrial inner membrane genes integral to the respiratory chain complex 1. Notably, two mitochondrial genes in these modules, *SLC25A4* (Pink_H), a component of the inner membrane, and *ATP5F1C* (Pink_G), an ATP synthase subunit, have been previously identified as *APOL1* binding partners that play a role in the mitochondrial permeability transition pore (mPTP) function^16^. Additionally, via CMAP, we identified *NDUFA8*, a component of respiratory chain complex I, as a potential upstream target differentiating HR vs LR FSGS. Previous work has shown that *APOL1* RVs activate opening of the mPTP which depolarizes the mitochondrial membrane leading to cytotoxicity^16^. Overall, the mitochondria appear to behave differently in the setting of HR vs LR gene expression, and some of the mitochondrial components of this response may contribute to driving the biology of APOL1 kidney injury.

Results from a number of analyses also pointed to differences in calcium regulation between HR and LR FSGS. *STC1* plays a role in calcium regulation, an observation that has been linked to mitochondrial function^56, 57^. The CMAP analysis of kidney cells found that a calcium channel agonist (synephrine) and inhibitor (levetiracetam) closely matched and opposed, respectively, the differential expression pattern in the HR vs LR group. Furthermore, it is recognized that levetiracetam has an impact on mitochondrial function^58-60^. Future analyses in relevant model systems are needed to directly determine whether calcium modulating drugs can alter *APOL1’s* cellular impact, the potential role of *STC1* in HR FSGS pathology vis-à-vis calcium dynamics, and the overall relationship between calcium regulation and mitochondrial function in glomerular cells.

The largest hub-gene in the entire analysis was NF-κB inhibitor interacting ras like 1 *(NKIRAS1). NKIRAS1’s* known function is to inhibit degradation of the NF-κB inhibitor beta (*NFKBIB*), which ultimately prevents NF-kB from traveling into the nucleus and initiation transcription^50 51^. There were no previous reports of *NKIRAS1* in relation to glomerular disease or *APOL1*. And while we did not clearly define its role in *APOL1-*associated FSGS in this study, we found it had strong differential co-expression with NDUF and JAK-STAT family genes. A subset of JAK-STAT genes showed strong negative correlations with *NKIRAS1* in the LR group but strong positive correlations in HR. JAK-STAT genes have previously been implicated in the pathology of FSGS and *APOL1* regulation^20^. NF-κB and the STAT genes also play a role in mitochondrial regulation^61 62^, raising interesting questions about potentially pleiotropic roles for these genes in *APOL1*-attributed FSGS and how *NKIRAS1* may fit in mechanistically. While the underlying biology remains to be discovered, our data suggests that the NF-κB pathway is differentially regulated in *APOL1*-associated FSGS.

There are a number of aspects to take into account when interpreting this study. First, mainly due to available sample sizes, we were agnostic to G1 vs G2 when classifying HR alleles and used the recessive model in all of our analyses, with 11 of 14 LR samples having one risk allele. In addition, because we compared two diseased groups, it is challenging to determine which signatures represent the potential contributory disease state. Using expression from normal glomerular tissue helped to discern the disease state. However, future experimental work in controlled, manipulable systems will be necessary to more precisely define disease signatures picked up in this study and more definitively dissect regulatory hierarchies and causal mechanisms.

In conclusion, this transcriptomic analysis of *APOL1*-associated FSGS in people extended mitochondrially-related discoveries from model systems, replicated immune-related findings from previous human analyses, and identified new genes and pathways for hypothesis-driven, mechanistic pursuit. Multiple lines of inquiry converged on the concepts that mitochondrial dysfunction, metabolic dysregulation, and a loss of gene coregulation in the HR state have a substantial and specific role in *APOL1*-associated FSGS. We highlighted the most impactful, cohesive findings from our analyses in the body of this paper, but there were many other significant relationships between *APOL1*, genes, and modules that we could not include here. Thus, we’ve created the “*APOL1* Portal” (http://APOL1portal.org) to make all of these analyses and all of the underlying expression data publicly available for browsing, download, and secondary analysis.

## Data Availability

We've created the APOL1 Portal (http://APOL1portal.org) to make all analyses in this paper and all of the data publicly available for browsing, download, and secondary analysis.

## Acknowledgments

The authors would like to thank Dr. Seongkyu Han and Dr. Christopher Benway for their discussions and contributions to Figures 1 and 3.

MS is supported by NIH RO1 DK108805, DK119380, and RC2 DK122397. DJ, NB, and JF are supported by 2 UM1 DK105554-06. The Nephrotic Syndrome Study Network Consortium (NEPTUNE), U54-DK-083912, is a part of the National Institutes of Health (NIH) Rare Disease Clinical Research Network (RDCRN), supported through a collaboration between the Office of Rare Diseases Research, National Center for Advancing Translational Sciences and the National Institute of Diabetes, Digestive, and Kidney Diseases. Additional funding and/or programmatic support for this project has also been provided by the University of Michigan, the NephCure Kidney International and the Halpin Foundation.

## Members of the Nephrotic Syndrome Study Network (NEPTUNE)

### NEPTUNE Enrolling Centers

*Cleveland Clinic, Cleveland, OH*: K Dell^*^, J Sedor^**^, M Schachere^#^, J Negrey^#^

*Children’s Hospital, Los Angeles, CA*: K Lemley^*^, B Silesky^#^

*Children’s Mercy Hospital, Kansas City, MO*: T Srivastava^*^, A Garrett^#^

*Cohen Children’s Hospital, New Hyde Park, NY:* C Sethna^*^, K Laurent ^#^

*Columbia University, New York, NY:* P Canetta^*^, A Pradhan^#^

*Emory University, Atlanta, GA:* L Greenbaum^*^, C Wang**, C Kang^#^

*Harbor-University of California Los Angeles Medical Center:* S Adler^*^, J LaPage^#^

*John H. Stroger Jr. Hospital of Cook County, Chicago, IL:* A Athavale^*^, M Itteera *Johns Hopkins Medicine, Baltimore, MD:* M Atkinson^*^, T Dell^#^

*Mayo Clinic, Rochester, MN:* F Fervenza^*^, M Hogan**, J Lieske^*^, V Chernitskiy^#^

*Montefiore Medical Center, Bronx, NY:* F Kaskel^*^, M Ross^*^, P Flynn^#^

*NIDDK Intramural, Bethesda MD:* J Kopp^*^, J Blake^#^

*New York University Medical Center, New York, NY:* H Trachtman^*^, O Zhdanova**, F Modersitzki^#^, S Vento^#^

*Stanford University, Stanford, CA:* R Lafayette^*^, K Mehta^#^

*Temple University, Philadelphia, PA:* C Gadegbeku^*^, S Quinn-Boyle^#^

*University Health Network Toronto:* M Hladunewich**, H Reich**, P Ling^#^, M Romano^#^

*University of Miami, Miami, FL:* A Fornoni^*^, C Bidot^#^

*University of Michigan, Ann Arbor, MI:* M Kretzler^*^, D Gipson*, A Williams^#^, C Klida^#^

*University of North Carolina, Chapel Hill, NC:* V Derebail^*^, K Gibson^*^, E Cole^#^, J Ormond-Foster^#^

*University of Pennsylvania, Philadelphia, PA:* L Holzman^*^, K Meyers**, K Kallem^#^, A Swenson^#^

*University of Texas Southwestern, Dallas, TX:* K Sambandam^*^, Z Wang^#^, M Rogers^#^

*University of Washington, Seattle, WA:* A Jefferson^*^, S Hingorani**, K Tuttle**^§^, M Bray^#^, E Pao^#^, A Cooper^#§^

*Wake Forest University Baptist Health, Winston-Salem, NC:* JJ Lin*, Stefanie Baker^#^

*Data Analysis and Coordinating Center*: M Kretzler*, L Barisoni**, J Bixler, H Desmond, S Eddy, D Fermin, C Gadegbeku**, B Gillespie**, D Gipson**, L Holzman**, V Kurtz, M Larkina, S Li, S Li, CC Lienczewski, J Liu, T Mainieri, L Mariani**, M Sampson**, J Sedor**, A Smith, A Williams, J Zee.

*Digital Pathology Committee*: Carmen Avila-Casado (University Health Network, Toronto), Serena Bagnasco (Johns Hopkins University), Joseph Gaut (Washington University in St Louis), Stephen Hewitt (National Cancer Institute), Jeff Hodgin (University of Michigan), Kevin Lemley (Children’s Hospital of Los Angeles), Laura Mariani (University of Michigan), Matthew Palmer (University of Pennsylvania), Avi Rosenberg (Johns Hopkins University), Virginie Royal (University of Montreal), David Thomas (University of Miami), Jarcy Zee (University of Pennsylvania) Co-Chairs: Laura Barisoni (Duke University) and Cynthia Nast (Cedar Sinai).

*Principal Investigator; **Co-investigator^; #^Study Coordinator

^§^Providence Medical Research Center, Spokane, WA

## Supplemental Methods

### Glomerular RNA-seq

Total RNA from 269 biopsies were prepared using the Clontech SMARTSeq v4 kit. Samples underwent sequencing using Illumina HiSeq 2500, resulting in 150bp unstranded, paired-end reads. Fastq files underwent quality control filtering and trimming using fastQC, fastQScreen^1^, and Picard Tools (http://broadinstitute.github.io/picard/). Trimmed reads were aligned to the human genome (GRCh37) with STAR 2.6.0a^2^. Gene expression counts were quantified using StringTie v2.1.4^3^. The genes were filtered to protein-coding genes with at least ten normalized counts in 14 samples. Variance stabilizing transformation was applied to counts in DESeq2^4^. The transformed reads were quantile normalized (https://github.com/bmbolstad/preprocessCore) and corrected for batch effects with Combat^5^. We used sampleNetwork^6^ to identify samples with outlying expression profiles, standardized connectivity < −3. This process was repeated until no outliers remained.

### Differential Coexpression

Differential co-expression gene modules were built using methods outlined by DiffCoEx^16^ (see Supplement for full details). We calculated the adjacency matrix of Spearman correlations for high-risk and low-risk separately. We then computed the matrix of adjacency differences, ranging from 0 to 1, where 1 indicates the strongest difference in correlations and 0 indicates no difference, followed by the topology overlap matrix (TOM) using the TOMdist function from WGCNA. The TOM was then clustered using flashClust with the “average” method. Modules were identified with the WGCNA function cutreeDynamic, with the “hybrid” method, cutHeight=0.996, deepSplit=T, pamRespectsDendro=F, and minClusterSize =20. Close modules were merged with mergeCloseModules using a cutHeight=0.2. Intra-modular connectivity for each gene was defined as the sum of adjacencies (from the matrix of adjacency differences) normalized by the largest connectivity value in the gene module; thus, the most connected gene would have normalized intra-modular connectivity of 1. Hub genes were defined as genes with intra-modular connectivity greater than 0.7. We used Cytoscape (v3.7.2) to visualize the correlation networks utilizing the ExpressionCorrelation (v1.1.0) app. Submodules were identified by comparing the genes’ hierarchical clustering within a DiffCoEx module in high-risk and low-risk separately. Permutation methods described by Tesson BM et al. ^16^ were used to test significance of gene modules. Submodule eigengenes (1^st^ principal component) were used to compare patterns between submodules.

---

**Supplemental Figure 1.**
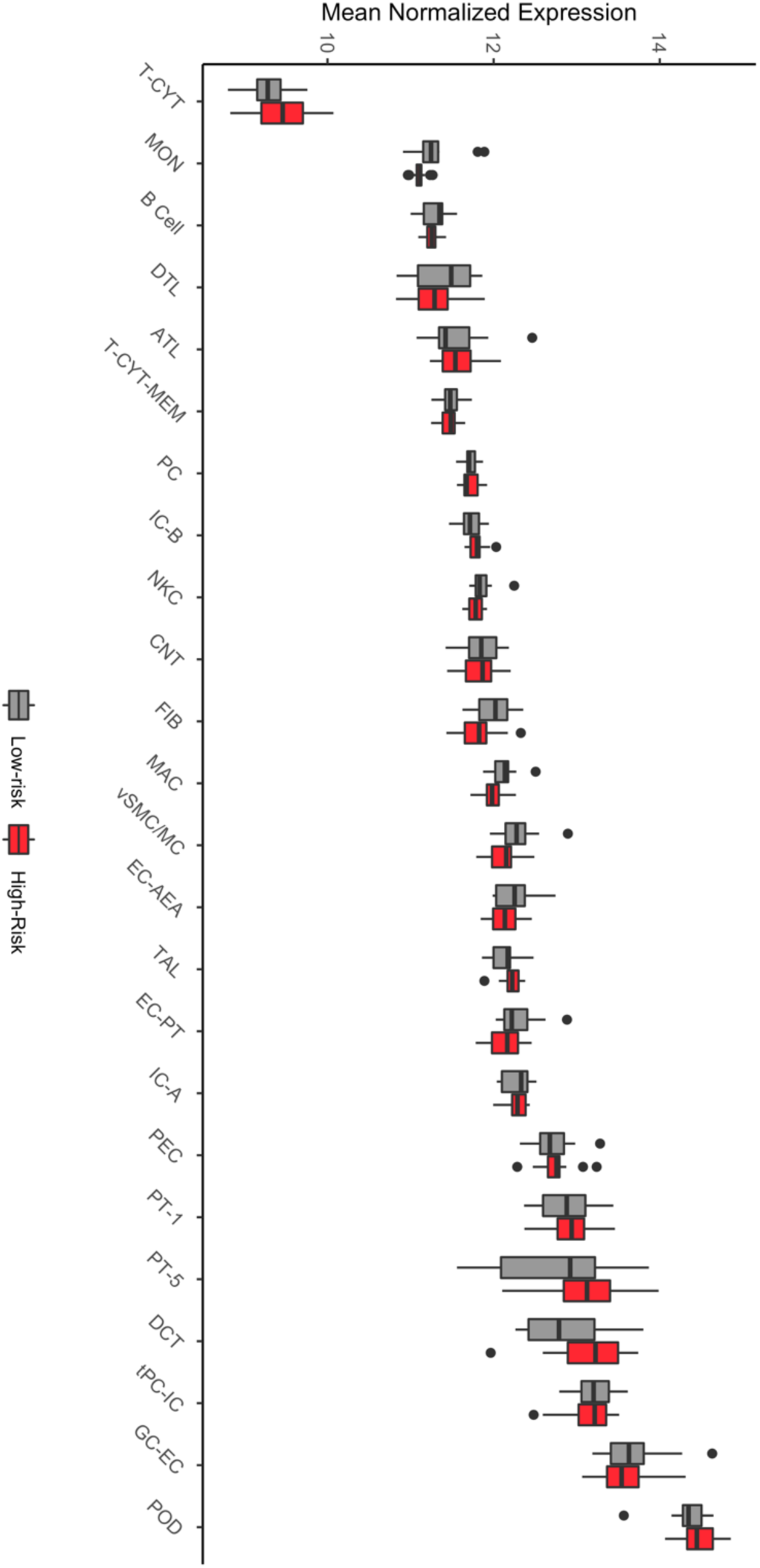
Mean expression of cell-type-specific genes per person stratified by high-risk and low-risk *APOL1* genotype: No significant differences were identified using a Bonferroni adjusted threshold = 0.004. T-CYT=cytotoxic T-cell, MON=monocyte, DTL=descending thin limb cell, ATL=ascending thin limb cell, T-CYT-MEM=memory T-cell, PC=principal cell, IC-B=intercalated beta cell, NKC=NK cells, CNT=connecting tubule, FIB=fibroblast, MAC=macrophage, vSMC/MC=vascular smooth muscle cell and mesangial, EC-AEA=endothelial cell, TAL=thin ascending limb cell, EC-PT=peritubular endothelial cell, IC-A=intercalated alpha cell, PEC=parietal epithelial cell/loop of Henle, PT-1=proximal tubule, PT-5=proximal tubule, DCT=distal convoluted tubule cell, tPC-IC=transitioning principal cell/intercalated cell, GC-EC=glomerular capillary endothelial cell, POD=podocyt

**Supplemental Figure 2.**
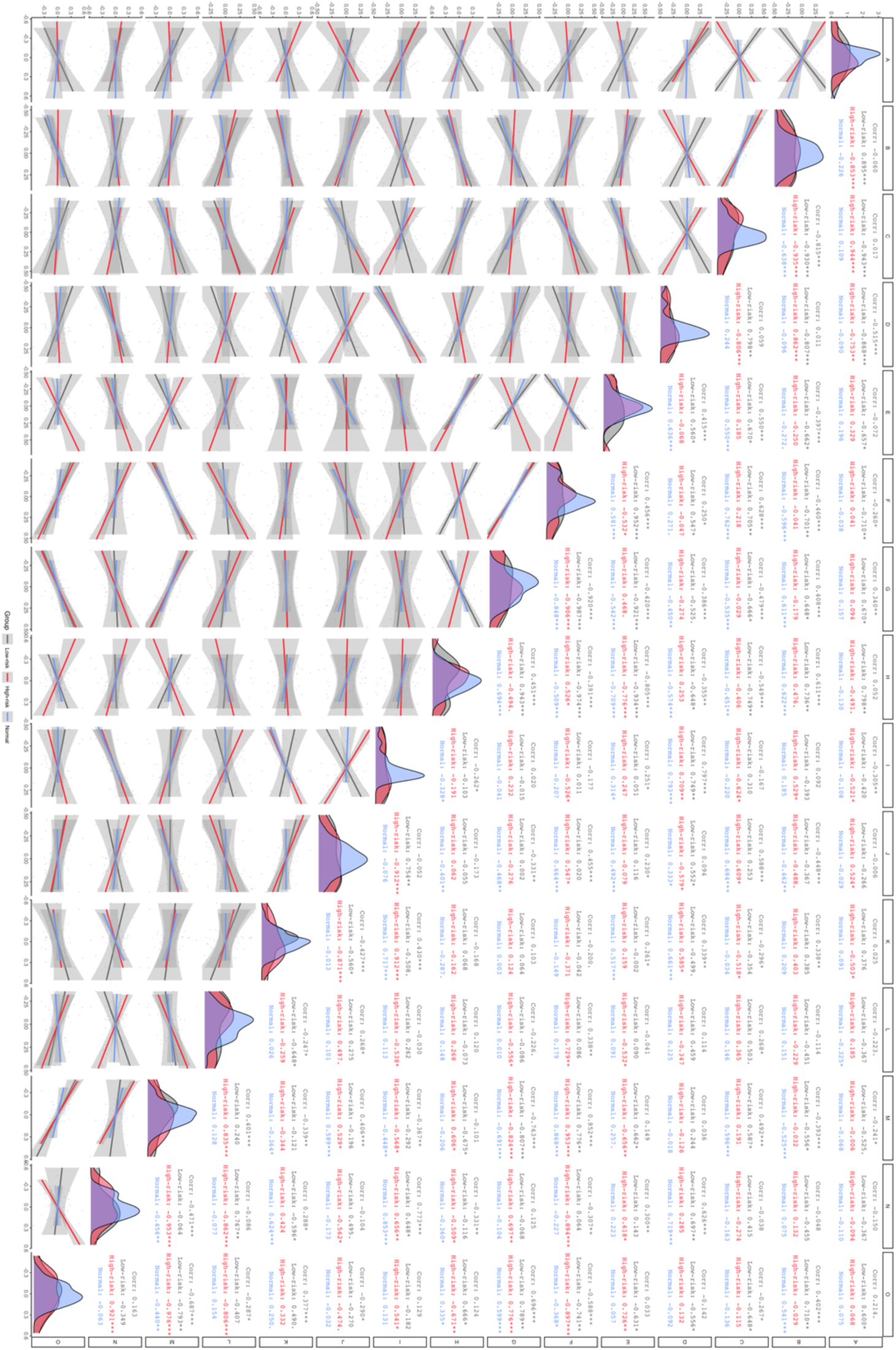
Eigengene Spearman correlations for all differentially co-expressed submodules in high-risk, low-risk, and healthy glomerular tissue: Eigengenes summarized gene expression behavior in each submodule and were defined as the first principal component of the group-specific gene expression matrices. Submodules are labeled along the x and y-axis. Diagonal – eigengene density plots. Below diagonal – scatter plot comparing submodule eigengenes (each point represents a sample). Above diagonal – Spearman correlations stratified by group (“Corr” is correlation in groups combined). Correlation significance - “***” p < 0.01, “**” p<0.01, “*” p<0.05, “.” p<0.

**Supplemental Figure 3.**
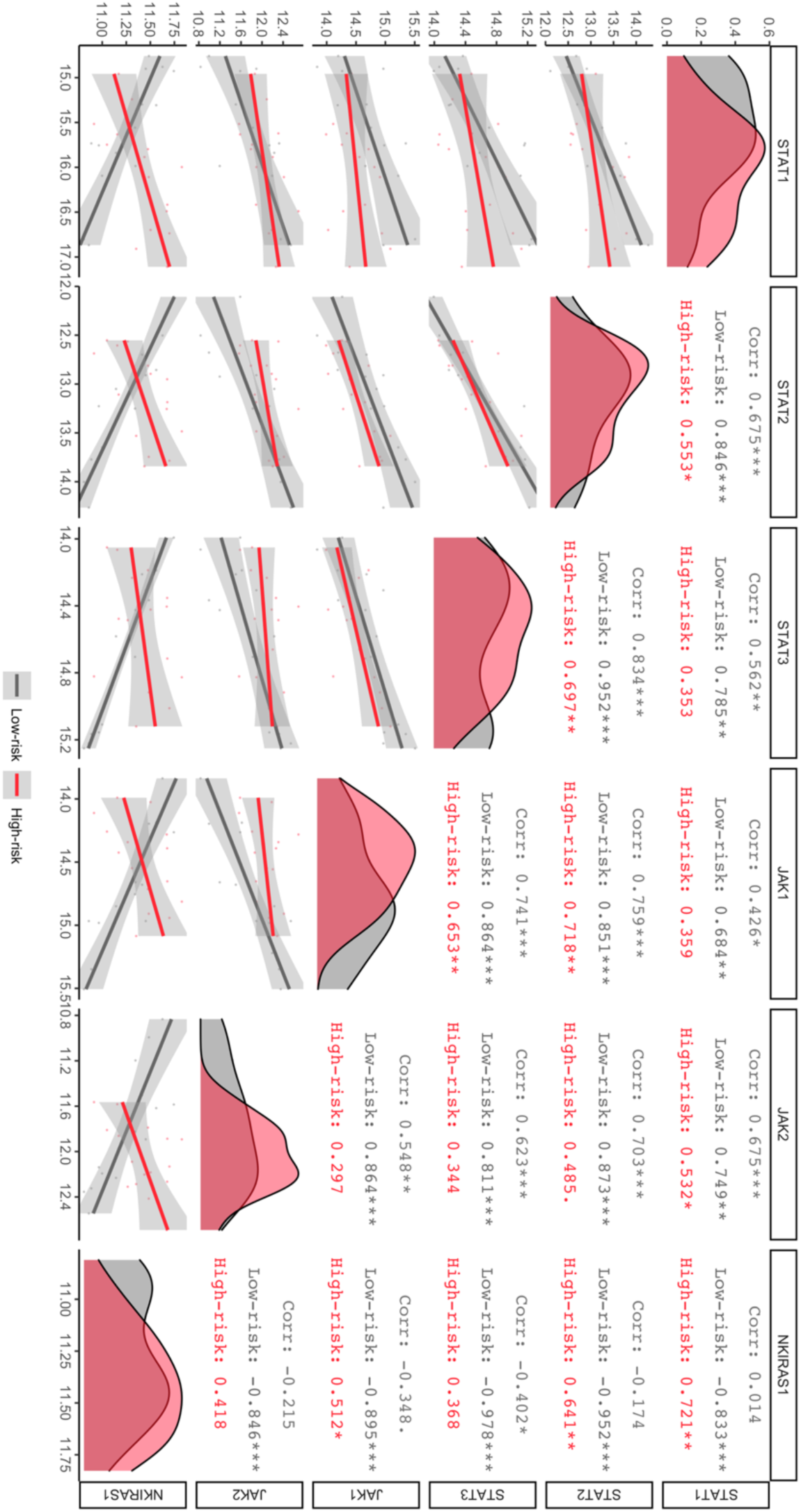
Spearman correlations between *JAK*/*STAT* genes from submodule E and hub-gene *NKIRAS1* stratified by high-risk and low-risk: Genes are labeled along the x and y-axis. Diagonal –gene expression density plots. Below diagonal – scatter plot comparing gene expression (each point represents a sample). Above diagonal – Spearman correlations stratified by group (“Corr” is correlation in groups combined). Correlation significance - “***” p < 0.01, “**” p<0.01, “*” p<0.05, “.” p<0.1

**Supplemental Figure 4.**
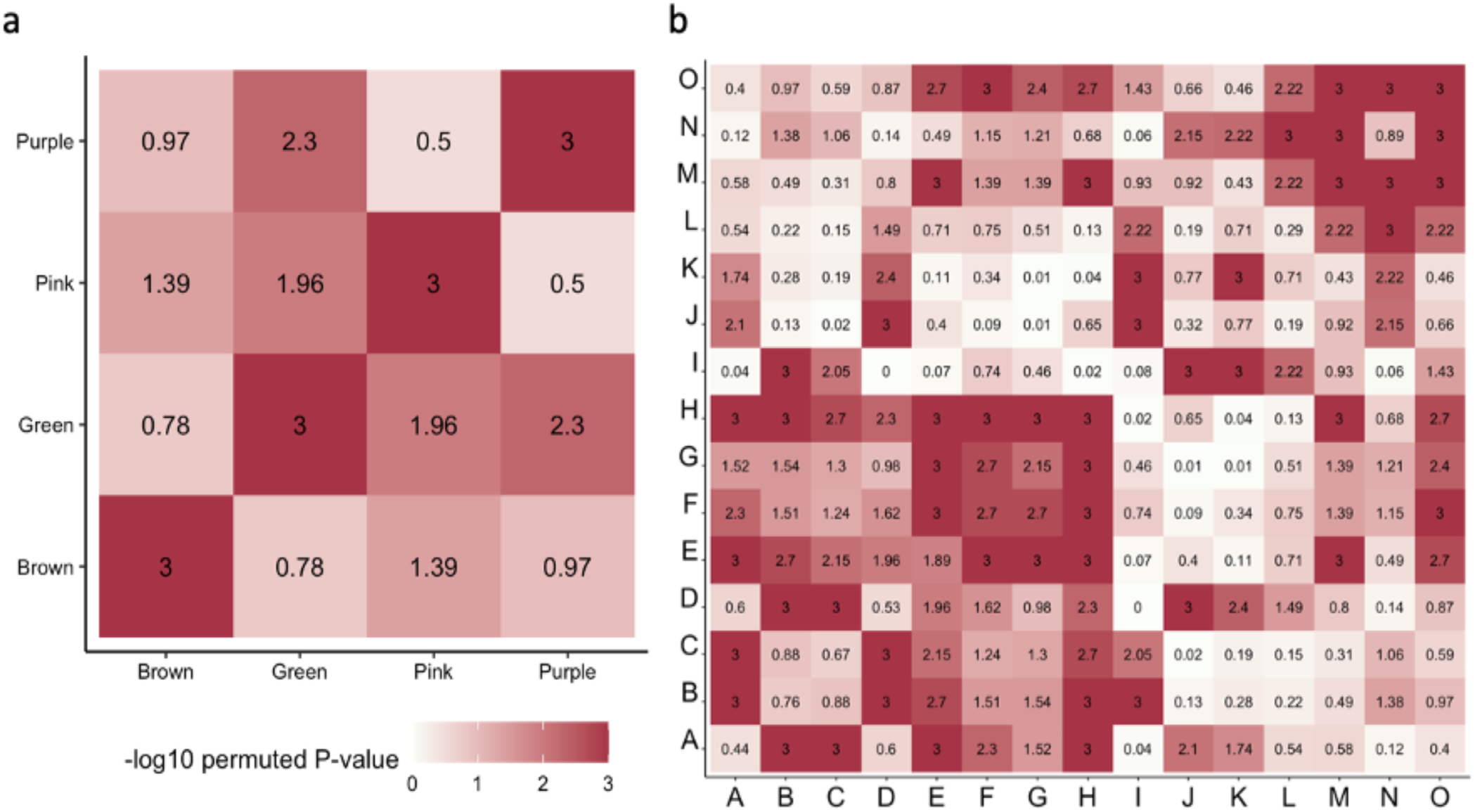
Significance of correlation changes within (diagonal) and between (off-diagonal) differentially co-expressed modules (A) and submodules (B): Both heatmaps are symmetric. For each figure, 1,000 permutations of samples were performed, and dispersion (correlation changes between groups) was calculated. We recorded the number of times the permuted data set resulted in more dispersion than our HR vs. LR analysis. We then -log10 transformed the permuted p-value (number of permutations with higher dispersion / 1,000). A -log10 p-value of 3 indicates changes in correlation patterns between HR and LR were stronger than any 1,000 permutation sample sets.

